# The Use of Electronic Health Record Embedded MRC-ICU as a Metric for Critical Care Pharmacist Workload

**DOI:** 10.1101/2023.09.27.23296158

**Authors:** Andrew J Webb, Bayleigh Carver, Sandra Rowe, Andrea Sikora

**Affiliations:** Oregon Health and Science University, Portland, OR; Massachusetts General Hospital, Boston, MA; University of Georgia College of Pharmacy, Department of Clinical and Administrative Pharmacy, 120 15th Street, HM-118, Augusta, GA

**Author notes:** **Correspondence:** Andrea Sikora Newsome, PharmD, MSCR, BCCCP, FCCM University of Georgia College of Pharmacy 120 15th Street, HM-118 Augusta, GA 30912.

**Keywords:** pharmacy practice models, critical care, medication regimen complexity, medication safety, clinical pharmacy, intensive care

## Abstract

**Objective:** A lack of pharmacist-specific risk-stratification scores in the electronic health record (EHR) may limit resource optimization. The medication regimen complexity-intensive care unit (MRC-ICU) score was implemented into our center’s EHR for use by clinical pharmacists. The purpose of this evaluation was to evaluate MRC-ICU as a predictor of pharmacist workload and to assess its potential as an additional dimension to traditional workload measures.

**Materials:** Data were abstracted from the EHR on adult ICU patients, including MRC-ICU scores and two traditional measures of pharmacist workload: numbers of medication orders verified and interventions logged.

**Methods:** This was a single-center study of an EHR-integrated MRC-ICU tool. The primary outcome was the association of MRC-ICU with institutional metrics of pharmacist workload. Associations were assessed using the initial 24-hour maximum MRC-ICU score’s Pearson’s correlation with overall admission workload and the day-to-day association using generalized linear mixed-effects modeling.

**Results:** A total of 1,205 patients over 5,083 patient-days were evaluated. Baseline MRC-ICU was correlated with both cumulative order volume (Spearman’s rho 0.41, p < 0.001) and cumulative interventions placed (Spearman’s rho 0.27, p < 0.001). A one-point increase in maximum daily MRC-ICU was associated with 31% increase in order volume (95% CI 24-38%) and 4% increase in interventions (95% CI 2-5%).

**Discussion:** The MRC-ICU is a validated score that has been previously correlated with important patient-centered outcomes. Here, MRC-ICU was modestly associated with two traditional objective measures of pharmacist workload, including orders verified and interventions placed, which is an important step for its use as a tool for resource utilization needs.

**Lay Summary:** Measuring critical care clinical pharmacist workload is challenging because currently available metrics, including number of medication orders verified or medication interventions logged, do not capture the full breadth of work critical care pharmacists do. The medication regimen complexity-intensive care unit (MRC-ICU) score is a tool designed to quantify the complexity of an ICU patient’s medication regimen and may serve as an alternative measure of overall critical care pharmacist workload. In this study, we assessed whether MRC-ICU scores from 1,205 ICU patients admitted to a single academic medical center were correlated with traditional metrics used to assess pharmacist workload, including medication orders and documented interventions. MRC-ICU was correlated with both workflow measures and traditional measures of patient acuity and also was predictive of the next day’s workload, suggesting MRC-ICU could be explored as an additional tool to optimize critical care pharmacist resource utilization. Further studies should assess how MRC-ICU can be utilized to optimize critical care pharmacist workload.

## BACKGROUND AND SIGNIFICANCE

Healthcare information technology (IT) tools designed for critical care pharmacists to streamline workload are not widely available.[1–3] However, workload has been related to patient outcomes for both intensivists and intensive care unit (ICU) nurses and remains a perennial challenge for critical care pharmacists.[4] As such, tools embedded within the electronic health record (EHR) that are capable of predicting the intensity or volume of patient care required from critical care pharmacists to help administrators make staffing decisions, and moreover, tools that can help make clinical workflow more efficient have the potential to improve patient outcomes.[5]

Current measures of pharmacist workload include elements such as volumes of medication orders verified and interventions logged. Advantages of such metrics include their readily quantifiable nature in the EHR and wide use among institutions; however, these have been repeatedly cited for their limitations in capturing the true workload of a critical care pharmacist.[2,5,6] The medication regimen complexity-ICU (MRC-ICU) scoring tool was designed as a measure that would correlate with the holistic care provided by critical care pharmacists. Moreover, its intent is to be an objective and reproducible metric to describe critical care pharmacists’ workload between diverse institutions. The score assigns a range of point values to a predefined list of medications and interventions stratified by the level of complex care required of a clinical pharmacist to manage that patient. For example, an order for an as-needed opioid analgesic would be assigned 1 point, while an order for a continuous opioid infusion would be assigned 2 points. All points are summed to calculate a patient’s MRC-ICU score. This tool has been validated and shown to correlate with key patient-centered outcomes (e.g., mortality, length of stay), complications of the Intensive Care Unit (ICU) stay (e.g., fluid overload, drug-drug interactions), and one measure of pharmacist workload (i.e., interventions).[6–11] Prior research, however, on the MRC-ICU score has been conducted using retrospective chart review to calculate scores and its real-time use as a tool in the EHR has not been previously described. The purpose of this study was to evaluate the ability of EHR-embedded MRC-ICU data to correlate with and predict traditional measures of pharmacist workload to assess whether MRC-ICU could serve as an additional dimension to commonly used objective metrics of pharmacist workload. Specifically, we hypothesized that MRC-ICU would show correlation with numbers of interventions logged and medication orders verified.

## METHODS

This was a single-center evaluation of an EHR-integrated MRC-ICU tool at Oregon Health and Science University (OHSU), a 576-bed academic medical center in Portland, OR. The study was evaluated and deemed exempt for review by the institutional review board (IRB). The pharmacy practice model at OHSU has clinical pharmacists integrated within medical teams that are responsible for rounding and clinical support as well as order verification and operational (i.e., medication dispensing) troubleshooting for the patients under their care. Pharmacists complete a full chart review of all patients each day and use electronic task lists for pharmacy consults and patient care follow ups. Unit-based clinical pharmacy coverage is available for 16 hours per day. The MRC-ICU tool was initially built into the EHR (Epic Systems, Verona, WI) as part of a multi-pronged approach to manage COVID-19 related patient surges to provide a means for pharmacists to prioritize review of high-acuity patients and remains available for use by clinical staff.[12] The MRC-ICU scoring tool can be found in **Table S1**. The use and evaluation of the score was left to the discretion of the pharmacists.

### Data Collection and Outcomes

Data were collected on all patients admitted to four specialty adult ICUs (medical, neurological/neurosurgical, trauma/surgical, and cardiovascular/cardiothoracic surgical) from November 1, 2020 to January 31, 2021. Collected data included basic patient demographics including age, gender, admitting unit, and discharge disposition, all MRC-ICU scores calculated during ICU admission which recalculated hourly, sequential organ failure assessment (SOFA) scores which recalculated once daily, number of medication orders placed and verified by pharmacists each day during ICU admission, and number of voluntarily logged pharmacist interventions (using the Epic i-Vent functionality). Interventions could include changes to patients’ medication plans including medication list optimization (e.g., changing a medication from intravenous to oral), patient safety measures (e.g., avoiding potential adverse drug events), or contributions to complex decision making (e.g., initiating therapy based on patient-specific factors). Interventions are a commonly recognized measure of pharmacist cognitive effort and are a potential metric of pharmacist workload in addition to cumulative verified orders verified.[13] Intervention documentation practices vary between pharmacists and it is unlikely that every pharmacist contribution to patient care is captured in a recorded i-Vent.

The primary objective of this study was to report the performance of data extracted from an EMR-embedded MRC-ICU as a predictor of commonly accepted objective measures of pharmacist workload, namely numbers of interventions logged and medication orders verified. Interventions and verified medication orders were selected as the measures of pharmacist workload because they are the most common metrics utilized by administrative departments to determine clinical pharmacist resource allocation and approval of additional clinical pharmacist positions. The MRC-ICU at baseline was used to assess correlation with overall admission workload, as was the correlation of daily MRC-ICU with daily workload and the performance of the MRC-ICU in predicting the next day’s workload. Secondary endpoints included evaluation of the correlation of MRC-ICU with SOFA scores, a commonly accepted measure of severity of illness, and the association with MRC-ICU and in-hospital mortality.

The MRC-ICU tool was available for pharmacist use but a specific approach to utilization was not mandated. Use of the tool included sorting patient lists by acuity to improve targeted patient workup (i.e. reviewing highest acuity patients first at the start of a shift), improved handoff (highlighting high acuity patients during handoff), and documentation (noting MRC-ICU scores in documentation). I-Vent documentation practices were also not standardized but general documentation were encouraged department wide; documentation included routine clinical/operational activities (IV to PO switches, route changes, renal dose adjustments) as well as more complex, involved decisions and clinical input (avoidance of unnecessary and/or potentially harmful therapies, addition of missing indicated therapies, rationale for specific therapeutic decisions made).

### Statistical Analysis

Baseline characteristics were reported as frequencies and percentages or medians and interquartile ranges (IQR) as appropriate. The maximum MRC-ICU score in the first 24 hours of a patient’s ICU admission has historically been the strongest predictor of patient outcomes in retrospective evaluations of MRC-ICU performance (i.e. calculating MRC-ICU on previously admitted patients) and was used as the baseline measure of complexity. Maximum MRC-ICU scores in each subsequent 24-hour period during ICU admission were also calculated and used to assess performance over time. Each individual patient’s total number of verified medication orders placed and interventions logged by pharmacists during ICU admission and daily were summed to represent the cumulative admission order volume and daily order volume. Because intervention documentation practices varied significantly between individual pharmacists, the number of recorded daily interventions was normalized across the range of daily interventions for each pharmacist using min-max feature scaling. That is, the minimum and maximum number of logged interventions in a day per pharmacist were scaled to fit the range of 0 to 1. Normalized interventions were also summed by patient day and cumulatively during a patient’s admission. This was completed to normalize cognitive effort across pharmacists. Because of documentation practice variation, the “weight” of an individual logged i-Vent cannot always be considered to be equal across pharmacists, meaning that an individual logged i-Vent from one pharmacist may not represent an equal unit of cognitive effort as a logged i-Vent from another pharmacist. The minimum and maximum number of intervention placed per patient, per day, per pharmacist was calculated to account for this. Because outliers in i-Vent logging is to be expected however, the analysis was also conducted using actual i-Vent counts.

The relationship between baseline maximum initial 24-hour MRC-ICU and cumulative admission normalized interventions and medication orders placed was assessed using Spearman’s correlation. Because MRC-ICU is a score mathematically derived from patient’s medication orders, MRC-ICU and number of medication orders are not fully independent and thus the correlation between maximum daily MRC-ICU and daily SOFA was assessed using Spearman’s correlation to evaluate whether MRC-ICU correlated with an alternative measure of patient acuity. Spearman’s rho was selected as a robust alternative to Pearson’s correlation as variables were not strictly linearly associated but were monotonic. Spearman’s rho is also less sensitive to outliers compared to Pearson’s correlation. Maximum initial 24-hour MRC-ICU was also assessed for an association with all-cause in-hospital mortality using logistic regression, adjusted for age, gender, and SOFA score.

To assess the performance of MRC-ICU over the course of a patient’s ICU admission, the relationship between daily maximum MRC-ICU and daily normalized interventions and medication orders was assessed using a generalized linear mixed-effects model with individual patients as a random effect. This was also conducted with SOFA scores. The ability of MRC-ICU to predict future workload was also assessed using the prior day’s maximum MRC-ICU score as a predictor for the next day’s medication orders and intervention volume using the same method. The association between baseline MRC-ICU and in-hospital mortality was conducted using logistic regression, adjusting for relevant baseline demographics (age, gender) as well as organ-based acuity using SOFA. Analysis was completed using R version 4.0.0 (R Core Team, R Foundation for Statistical Computing, Vienna, Austria) with linear mixed effects models being constructed using the *lme4* package.[14]

## RESULTS

A total of 1,242 patients over a three-month period equating to 5,083 patient-ICU days were evaluated resulting in 42,482 medication orders verified, 3,394 i-Vents placed, and 78,515 scores generated. After min-max feature scaling, the total number of normalized i-Vents placed was 580 (average ratio of 5.85:1 total to normalized i-Vents). Baseline demographics are summarized in **Table 1**. Patients had a median age of 62 years, had a median hospital length of stay of 8 days, and most were discharged home from the hospital (65%). Using coded admission ICD-10 codes, 2.3% of admission diagnosis codes were listed as COVID-19 while 7.2% of admission codes pertained to respiratory failure or acute respiratory distress syndrome (ARDS). Overall, 8.8% of patients died during their admission and 5.8% were discharged to hospice.

**Table 1:** Baseline Demographics.

| Characteristic | N = 1,242<sup>1</sup> |
| --- | --- |
| Age, years | 62 (48, 72) |
| Gender |  |
| Female | 500 (40%) |
| Male | 742 (60%) |
| Admitting ICU |  |
| CTICU | 452 (36%) |
| TSICU | 364 (29%) |
| NCCU | 270 (22%) |
| MICU | 156 (13%) |
| Hospital Length of Stay, Days | 8 (4, 15) |
| Discharge Disposition |  |
| Home | 796 (65%) |
| SNF | 164 (13%) |
| Deceased | 107 (8.8%) |
| Hospice | 71 (5.8%) |
| Inpatient Rehab | 31 (2.5%) |
| LTCH | 20 (1.6%) |
| Inpatient Transfer | 18 (1.5%) |
| Against Medical Advice | 11 (0.9%) |
| Total ICU Medication Orders |  |
| CTICU | 13,410 (31.6%) |
| TSICU | 10,541 (24.8%) |
| NCCU | 8,808 (20.7%) |
| MICU | 9,723 (22.9%) |
| Total ICU i-Vents |  |
| CTICU | 977 (28.8%) |
| TSICU | 1,152 (33.9%) |
| NCCU | 616 (18.1%) |
| MICU | 649 (19.1%) |
| Total Normalized ICU i-Vents |  |
| CTICU | 162 (27.9%) |
| TSICU | 167 (28.8%) |
| NCCU | 118 (20.3%) |
| MICU | 133 (22.9%) |

<sup>1</sup>Median (IQR); n (%); Range
CTICU, cardiothoracic intensive care unit; LTCH, long-term care hospital; MICU, medical intensive care unit; NCCU, neurocritical care unit; SNF, skilled nursing facility; TSICU, trauma/surgical intensive care unit

The maximum initial 24-hour MRC-ICU correlated with both cumulative ICU admission medication orders (Spearman’s rho 0.41, p < 0.001) and cumulative normalized i-Vents (Spearman’s rho 0.27, p < 0.001). MRC-ICU at 24 hours was also correlated with admission SOFA score (Spearman’s rho 0.31, p < 0.001). When evaluating the relationship between daily maximum MRC-ICU and medication orders placed, each 1-point increase in daily maximum MRC-ICU was associated with an approximately 31% increase in daily medication orders (95% CI 24-38%) and 4% increase in normalized i-Vents (95% CI 2-5%). When the analysis was repeated using actual i-Vent counts instead of normalized i-Vent counts, results were similar (**Table S2**). The effect of MRC-ICU on order volume varied minimally across patients, with a mean random effect per patient of 0 and standard deviation of 0.057. The random effects of patients did not significantly contribute to the i-Vents model. Daily SOFA scores were not well-modeled by changes in daily maximum MRC-ICU; each 1-point increase in MRC-ICU was associated with an approximately 3% increase in daily SOFA score (95% CI 3-3%) but model variance between patients was large. The predicted values of daily medication orders, normalized i-Vents, and SOFA score across the range of MRC-ICU scores for each is depicted in **Figure 1**. To simulate the potential role of MRC-ICU as a next-day planning metric, the same models were constructed using the previous day’s maximum MRC-ICU for each patient. Each model performed similarly; a 1-point increase in the maximum daily MRC-ICU was associated with an 18% increase in the next day’s daily medication orders (95% CI 12-24%) and 3% increase in the next day’s daily normalized i-Vents (95% CI 1-5%). Model coefficients are listed in **Table S2.**

**Figure 1:**
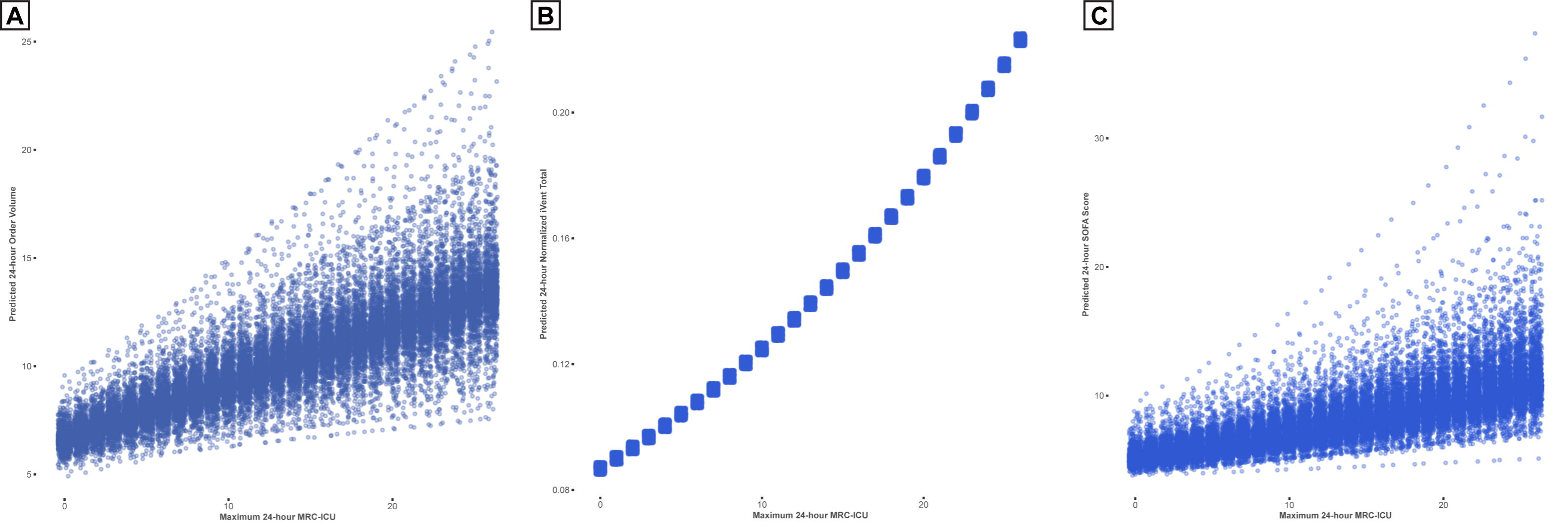
Predicted Values Based on Maximum Daily MRC-ICU: Predicted values of the generalized linear mixed effects models for daily medication orders, normalized i-Vents, and daily SOFA scores based on daily maximum MRC-ICU. Individual points represent predicted values per individual patient random effects. A: Medication Orders, B: Normalized i-Vents, C: SOFA

Maximum initial 24-hour MRC-ICU was also assessed for an association with all-cause in-hospital mortality (**Figure 2**). Each 1-point increase in baseline MRC-ICU was associated with an increase in the odds of in-hospital mortality (OR 1.14, 95% CI 1.09-1.20). Each 10-year increase in age (OR 1.16, 95% CI 1.02-1.33) and each 1-point increase in SOFA (OR 1.10, 95% CI 1.03-1.17) were also associated with increased odds of mortality while gender (OR 0.96, 95% CI 0.64-1.47) was not.

**Figure 2:**
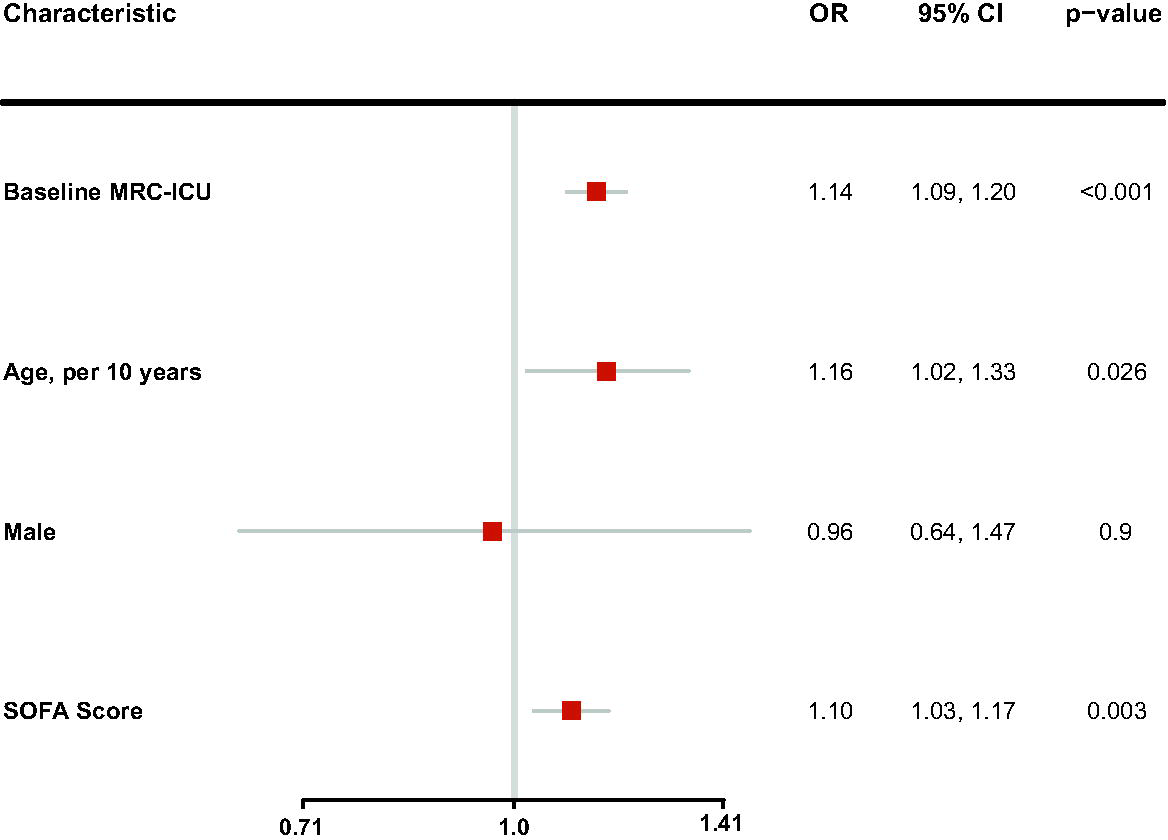
Factors Associated with In-Hospital Mortality: Multivariable model of factors associated with in-hospital mortality. Odds of in-hospital mortality are per 1-point increase in baseline MRC-ICU, 10-year increase in gender, male gender vs female gender, and 1-point increase in admission SOFA score. *MRC-ICU: medication regimen complexity, intensive care unit*; *OR: odds ratio; CI: confidence interval; SOFA: sequential organ failure assessment*

## DISCUSSION

In the first analysis of an integrated MRC-ICU into the EHR, MRC-ICU demonstrated correlation with two institutionally established metrics of pharmacist activity. We revealed that initial maximum admission MRC-ICU derived from the EHR was modestly correlated with cumulative workload volume over the entire course of ICU patients’ admission and that dynamic changes in the MRC-ICU day-to-day were associated with increases in daily medication order and pharmacist verification volume. These findings suggest that an EHR-based MRC-ICU report could be used as an additional descriptive tool to characterize pharmacist workload volume and may be helpful in resource utilization planning to dynamically allocate pharmacist resources to areas who may be expected to have high clinical pharmacist need and as a prioritization tool for those clinicians at the bedside.

MRC-ICU demonstrated modest correlation with interventions, medication orders verified, and SOFA score. In a previous evaluation, MRC-ICU showed correlation to number of medications and number of medication orders via Pearson coefficients of 0.4876 and 0.5308, respectively (p < 0.0001), which resulted in a thoughtful commentary discussing the trap of overvaluing a statistically significant p-value in the face of relatively modest correlation.[8,15,16] The authors’ reply noted that while the MRC-ICU is intended to objectively and consistently indicate the degree of medication regimen complexity as faced by clinicians managing patients in the ICU (and that this is related to overall number of medications ordered for a patient) that “the specific identity of the medication adds another layer of complexity beyond simply order volume. Indeed, if the score correlated too strongly with number of medications or number of medication orders, then this correlation could be argued to negate the need to develop the MRC-ICU altogether.” Similarly here, the goal of capturing medication regimen complexity is to reflect the cognitive workload of a clinician in evaluation of this regimen; such cognition may or may not always result in a discrete intervention. Ultimately, a metric like this would be well served for evaluation in relation to total time related to direct patient care or multiple metrics of pharmacist workload beyond interventions.

Despite being essential members of the interprofessional healthcare team, the optimal critical care pharmacy practice model and the associated critical care pharmacist to patient ratio remain unknown.[2,17] These knowledge gaps have resulted in notable limitations in the care that can be provided to patients in the ICU. Indeed, it has been reported that only 70% of ICUs have rounding pharmacists and this was mostly limited to weekday, day-time coverage.[18] Furthermore, burn-out syndrome is strikingly common in critical care pharmacy and increased patient care responsibilities have been associated with the development of burn-out.[19,20]

The EHR has been used in a number of innovative ways to measure and quantify clinical pharmacist activity, but workload measurement generally revolves around discrete, pre-defined activities that are easily documented in the EHR. This may include electronic task lists for pharmacist-driven protocol to-do items,[21] documenting the depth of daily patient chart review,[22] or development of institution-specific scoring tools for complex medication regimens.[23] These tools allow for institutional tracking of pharmacist activities, but often, they do not correlate with pharmacist workload or patient severity of illness. Notably, excess clinical activities not adequately captured may contribute to pharmacist burnout.[20] We demonstrated that MRC-ICU metrics could be used both as a marker of pharmacist workload, correlating both with medication orders verified and interventions logged, and patient acuity, correlating with SOFA scores and associated with in-hospital mortality. These relationships are in line with previous evaluations of the MRC-ICU, which have shown relationships to severity of illness scores, mortality, and workload.[6] The predictive nature of the score (i.e., associated with increasing workload the next day) provides a unique opportunity to utilize the EHR to evaluate potential clinical pharmacist needs the next day, potentially allowing administration to objectively identify when additional resources should be allocated to high-acuity areas.

Intervention tracking and cost avoidance for measuring pharmacist workload has numerous limitations.[5,24] A clinical pharmacist’s day includes both direct and indirect patient care activities and an as of yet unmeasured portion of the day goes toward ‘medication-related dialogue’ in which the pharmacist is participating as an active member of the interprofessional healthcare team to proactively create and implement treatment plans. Further, the critical care pharmacist’s value to the team goes well beyond verifying medication orders.^21^ Yet, both medication orders verified and interventions documented are readily quantifiable metrics that have historical importance in characterizing pharmacist activity and moreover are institutionally relevant.

Two key advantages of the MRC-ICU are the clinician-centered design and external validation efforts. The MRC-ICU was designed by clinicians to capture holistic care, which ultimately supports key human-machine teaming principle and future bedside adoption. Second, the external validation studies support generalizability for other institutions.[8] One institution developed a productivity model which sought to quantify clinical pharmacist work output beyond order verification alone, including unit census, number of “time intensive” medications, order verification volume, and transition of care activities. While certain elements of productivity were calculated beyond the simple quantification of medication orders verified, cognitive work (e.g., decision making during patient care rounds, development of clinical practice guidelines and protocols, educating learners, and conducting research) is difficult to quantify. As such, this score does not capture all elements of pharmacist output and would require additional tracking or measurement.[25,26] Moreover, such a model may be too institutionally specific for generalizability. As previously outlined by this team,[2,17], the process for developing holistic, clinically-relevant metrics that can foster inter-institution evaluation of best practices (such as following a step-wise approach and being able to predict patient care resources) is foundational to this process.

Limitations of this study include its small, single-center, non-interventional design which may limit external generalizability and inferential conclusions. While this study supports the feasibility of building an electronic MRC-ICU as a component of pharmacist workflow and utilizing MRC-ICU data from the EHR for workload assessment, it does not have the ability to assess impact of adjusting clinical pharmacist resources based on MRC-ICU on patient outcomes, which will need further, prospective analysis. The use of MRC-ICU as an administrative metric is purely exploratory in this context and should be evaluated in future investigation.

### Conclusion

An automated MRC-ICU may be integrated into the electronic health record for use by critical care pharmacists and may be able to be utilized as a predictive measure of future clinical staffing needs. Additional prospective study should be undertaken to better characterize how it correlates with additional facets of pharmacist workload as well as in additional clinical practice settings, especially at centers with higher patient workload per pharmacist.

## Data Availability

All data produced in the present study are available upon reasonable request to the authors.

## Funding Statement

This work was supported by AHRQ grant #1R01HS029009.

## Competing Interests

The authors have no competing interests to declare. This work was partially presented as a platform presentation at Epic User’s Group Meeting (Verona, WI) in August 2021.

## Contributorship Statement

AJW, SR, and AS were involved in project ideation and design. AJW and SR were involved in data collection. AJW was involved in data analysis. AJW, BC, SR, and AS were involved in data interpretation. BC wrote the initial manuscript draft, AJW, BC, SR, and AS were involved in critical manuscript edits.

## Data Availability

The data underlying this article will be shared on reasonable request to the corresponding author.

## Notes

### Competing Interest Statement

The authors have declared no competing interest.

### Author Declarations

IRB of OSHU waived ethical approval for this work.

