## Supplemental for "The Use of Electronic Health Record Embedded MRC-ICU as a Metric for Critical Care Pharmacist Workload"

**Table S1: MRC-ICU Scoring Tool**

Each individual medication or medication class listed is associated with a weighted score (i.e., 1, 2, or 3 points). If a multiplier is listed (e.g. 3x), this indicates that for every medication that meets those criteria, the score is multiplied (e.g., if fentanyl and midazolam are prescribed, this would be 2 points x each medication for a total of 4 points). The MRC-ICU is the sum of all the points assigned to the medications.

| High Priority Medications | Point Value |
| --- | --- |
| Aminoglycoside antibiotic | 3x |
| Amphotericin B Products | 1 |
| Antiarrhythmic Medications | 1x |
| Anticoagulants (DOAC, fondaparinux) | 1x |
| Anticonvulsants | 3x |
| Argatroban | 2 |
| Azole antifungals | 2x |
| Blood products | 2x |
| Inpatient chemotherapy | 3x |
| Clozapine | 3 |
| Digoxin | 3 |
| Ganciclovir/valganciclovir | 1x |
| Hyperosmolar fluid/product | 1x |
| Immunosuppressant | 3x |
| Continuous lidocaine | 2 |
| Lithium | 3 |
| Prostacyclin | 2x |
| Therapeutic heparin product | 2x |
| Vancomycin IV | 3 |
| Warfarin | 3 |
| ICU Medications |  |
| Neuromuscular blockade | 2 |
| Continuous infusion not otherwise specified | 1x |
| Total parenteral nutrition |  |
| Non-pharmacist managed | 1 |
| Pharmacist managed | 3 |
| ICU Prophylaxis |  |
| Thromboembolism prophylaxis | 1 |
| Stress ulcer prophylaxis | 1 |
| Glycemic control (excluding IV infusion) | 1 |
| Bowel regimen | 1 |
| Chlorhexidine | 1 |
| Analgesia and Sedation |  |
| Opioids and sedatives (scheduled and PRN) | 1x |
| Continuous infusion opioid or sedative | 2x |
| Antimicrobials |  |
| Antimicrobials not listed elsewhere | 1x |
| Restricted antimicrobial | 2x |
| Devices |  |
| Dialysis | 2 |
| ECMO | 2 |
| Intra-aortic balloon pump | 1 |
| LVAD | 1 |
| Mechanical ventilation | 2 |

*DOAC: direct oral anticoagulant; ECMO: extracorporeal membrane oxygenation; LVAD: left ventricular assist device; PRN: as needed*

**Table S2: Generalized Linear Mixed Effects Model Coefficients**

| Predicted outcome (per 1 unit increase in MRC-ICU) | Intercept | Coefficient (Type) | 95% CI | Random Effects (Per Patient) Variance | Random Effects SD |
| --- | --- | --- | --- | --- | --- |
| Orders | 6.62 | 0.27 (β) | 0.22-0.33 | 2.00 | 1.59 |
| Normalized i-Vents | 0.09* | 0.04 (logIRR) | 0.02-0.05 | <0.001 | <0.001 |
| Actual i-Vents | 0.40* | 0.04  (logIRR) | 0.03-0.05 | 0.25 | 0.50 |
| SOFA | 5.15* | 0.03 (logIRR) | 0.03-0.03 | 0.04 | 0.21 |
| Per 1 Point Increase in Prior Day MRC-ICU | | | | | |
| Orders | 6.09 | 0.17 (β) | 0.12-0.22 | 2.24 | 1.637 |
| Normalized i-Vents | 0.08* | 0.03 (logIRR) | 0.01-0.05 | <0.001 | <0.001 |
| Actual i-Vents | 0.39* | 0.03  (logIRR) | 0.02-0.04 | 0.24 | 0.49 |

β coefficients represent Gaussian (linear) regression coefficients; each 1-point increase in MRC is associated with a linear increase in the β coefficient. Log incidence rate ratio (logIRR, Poisson distribution) represents the change in the logarithm of the expected count for a one-unit change in MRC-ICU. In other words, each one unit increase in MRC-ICU increases the natural logarithm of the outcome by the coefficient.

*Intercept has been exponentiated for easier interpretation. True model intercepts can be calculated by taking the natural logarithm of the listed intercept.
